# The profile of urban healthcare service provision with a focus on maternal and newborn health in Grand Conakry, Guinea: Results of a 2025 health facility census

**DOI:** 10.64898/2026.08.19.26360676

**Authors:** Nafissatou Dioubate, Aline Semaan, Rehema Ouko, Gloria Jules Djidonou, Mohamed Koulako Keita, Pépé Kpogomou, Makany Sangare, Hadja Fatoumata Souare, Fassou Mathias Grovogui, Tamba Mina Millimouno, Hawa Manet, Peter Macharia, Souleymane Diakite, Abdoul Karim Nabe, Dieney Fadima Kaba, Facely Camara, Fodé Badara Conte, Jean Dobo Sovogui, Emmanuel Goumou, Abdoulaye Sano, Housseynatou Sow, Bansoumane Sougoule, Fanta Dabo, Aïssiatou Balde, Aminata Balde, Sékou Diafodé Sano, Saran Camara, Fatoumata Sanoh, Jean Pierre Leno, Madina Sissoko, Albert Soko Sagno, Bintou Conde, Alseny Camara, Sekou Niouma Camara, Mamadou Baïllo Bah, Sidikiba Sidibe, Alexandre Delamou, Lenka Beňová

## Abstract

**Background:** Rapidly urbanising complex ecosystems present several challenges to the organisation, availability, and accessibility of healthcare services. This study describes the availability and distribution of health facilities offering services, with a focus on maternal and newborn health services in the seven health districts of Grand Conakry,

**Methods:** This study using primary data from a census of health facilities (June 2025). Finally, healthcare landscape was drawn up using the GPS coordinates collected, based on the boundaries of the seven including health districts. The census started with an existing list of 459 facilities and snowball sampling was used to identify facilities that were not initially on the list. We collected information on basic facility characteristics and GPS coordinates.

**Results:** A total of 795 health facilities exist in Grand Conakry, with 564 facilities which were not on the initial lists, 104 were either closed, out of service, or refused to participate. The questionnaire was completed by 691 health facilities. The private sector dominated the landscape and was mainly concentrated in the Ratoma and Matoto health districts; with most facilities (63%) not reporting to the national health information system, in contrast to those in public sector (85.7%). 332/691 facilities reported providing childbirth care, and 132 of them (40%) offered caesarean section, while 87 (26%) offered blood transfusions.

**Conclusion:** Despite the high number of facilities, geographic imbalances in distribution exist. Equitable access to healthcare in rapidly urbanising Grand Conakry requires strengthening urban health planning and public-private collaboration regarding ongoing rapid urbanisation in Grand Conakry.

**Key messages:** *What is already known on this topic?:* The rapid urbanisation of Grand Conakry is accompanied by an expansion in healthcare provision, but information on the geographical distribution and characteristics of healthcare facilities, particularly for maternal and neonatal care is limited and incomplete.

*What this study adds?:* A significant number of healthcare facilities operating in Grand Conakry are missing from existing health authority lists. Healthcare provision is largely dominated by the private sector. The availability of comprehensive emergency obstetric and neonatal care services remains limited, and only 4% of facilities offer childbirth care that is completely free of charge (all public).

*How this study might affect research, practice or policy?:* The comprehensive facility list generated by this census strengthens healthcare system planning for maternal and neonatal care provision, including improving referral networks in Grand Conakry.

## INTRODUCTION

Rapid urbanisation in sub-Saharan Africa poses major challenges for the provision and accessibility of health services.^1^ This is mainly due to urban ecosystems’ complexity, resulting from dynamic interactions between demographic, economic, environmental and organisational factors. Rapid population growth leads to the expansion of informal settlements and the predominance of the informal economic sector characterised by irregular income sources for most of the working population.^1–3^ Additionally, the occupation of ecologically vulnerable areas exposed to the risk of natural disasters (flooding, erosion) increases the risk of infectious diseases and the barriers to accessing health services.^4^ Furthermore, this urban environment combines household structural economic vulnerability with insufficient healthcare financing, thereby exacerbating health disparities and revealing a gap in the health system’s capacity to adapt to the accelerated pace of urbanisation.^5,6^ Therefore, adequately assessing and managing health needs in urban settings is essential to promoting the sustainable health and well-being of the population.

Urbanisation can offer advantages in terms of healthcare provision and utilisation, notably in health infrastructure and health staff concentration, development of road infrastructure or better reach of health information and innovations.^7,8^ However, in some sub-Saharan African cities, evidence indicates that the urban health advantage is declining.^9^ A study in two slums in Lagos (Nigeria, 2017) revealed a maternal mortality ratio (MMR) of 1,050 deaths per 100,000 livebirths compared to 545 per 100,000 livebirths in Lagos State.^10^ Similarly in Nairobi (Kenya, 2003-2005), disadvantaged urban areas had a MMR of 706 deaths per 100,000 livebirths, compared to the national level (560 per 100,000).^11^ In Tanzania, a study showed that core urban areas have perinatal mortality twice as high compared to rural clusters.^9^

For several decades since independence from French colonisation in 1958, the Grand Conakry metropolitan area in Guinea underwent rapid development accompanied by sustained population growth. The term “Grand Conakry” remains relatively recent in Guinea’s political and urban vocabulary, emerging in the 2010s as part of a vision to expand and modernize the capital.^12^ This initiative involved integrating peri-urban areas of surrounding towns in Coyah and Dubréka districts into a unified urban development plan of Conakry. Consequently, historical population estimates for Grand Conakry are limited, unlike those for the Conakry administrative region,^13,14^ where population estimates were 112,000 in 1960, 1.5 million in 2010, and 2.25 million 2026.^15,16^

The number of health facilities also increased significantly over time, notably through the development of primary care facilities (public health centres and health posts), second level of care (two prefectural hospitals, four district medical centres), and the private sector (clinics, medical practices or polyclinics).^3,11^ However, significant disparities exist in the distribution of facilities within Grand Conakry.^17,18^ During the early post-independence period, healthcare services were concentrated in the city centre (the wealthier neighbourhood’s) and limited to few public modern facilities, such as Ignace Deen and Donka hospitals, supplemented by health posts and basic clinics. With urbanisation and other developments since 1990, the concentration of health facilities shifted to the outskirts. Several historical large public hospitals remain in the city centre, which now hosts administrative and economic activities rather than residential areas. To meet the growing demand for healthcare, new health facilities (mostly private sector) were built in peripheral and suburban areas, where the middle-class and wealthier populations now reside. Meanwhile, informal settlements and peri-urban areas remain underserved.^19^ Additionally, many health facilities, particularly national hospitals, have not significantly increased their capacity since establishment,^1,20,21^ and are thus unable to respond to rapidly growing demand for advanced and emergency care.

It is essential to update health service availability to respond to the changing needs. Previous studies of Grand Conakry used secondary data on healthcare availability, which may be incomplete or insufficiently updated, especially with regards to the private sector.^5,19^ Also, information on services offered by facilities is not always directly available, but inferred (from level) or imputed from other source, which may limit accuracy.^22^ It is therefore urgent to update and comprehensively map information on health facilities to accurately describe the availability and affordability of maternal healthcare services. The objective of this study is to map and analyse the availability and distribution of health facilities in Grand Conakry, with a focus on maternal and newborn health services.

## METHODS

### Study setting

This study was conducted in Grand Conakry, a metropolitan area encompassing the central urban area of Conakry city Conakry and the peri-urban areas of Coyah and Dubréka (Figure 1). Grand Conakry comprises five health districts in Conakry (Kaloum, Matam, Dixinn, Ratoma and Matoto), one in Dubréka and one in Coyah.^19^ Since the 1960s, Guinea has experienced sustained urbanisation. The proportion of the population living in urban areas rose from around 10% in 1960 to 37% in 2022. Conakry accounts for more than half (54%) of the country’s urban population. In 2024, the population of Grand Conakry was estimated at over 2.6 million, of whom 49.5% are women. Significant proportions of this population face socioeconomic vulnerabilities, with 43.7% living below the poverty line.^23^ In 2018, the national total fertility rate was 3.8 births per woman in urban areas compared to 5.5 in rural areas.^24^

**Figure 1.**
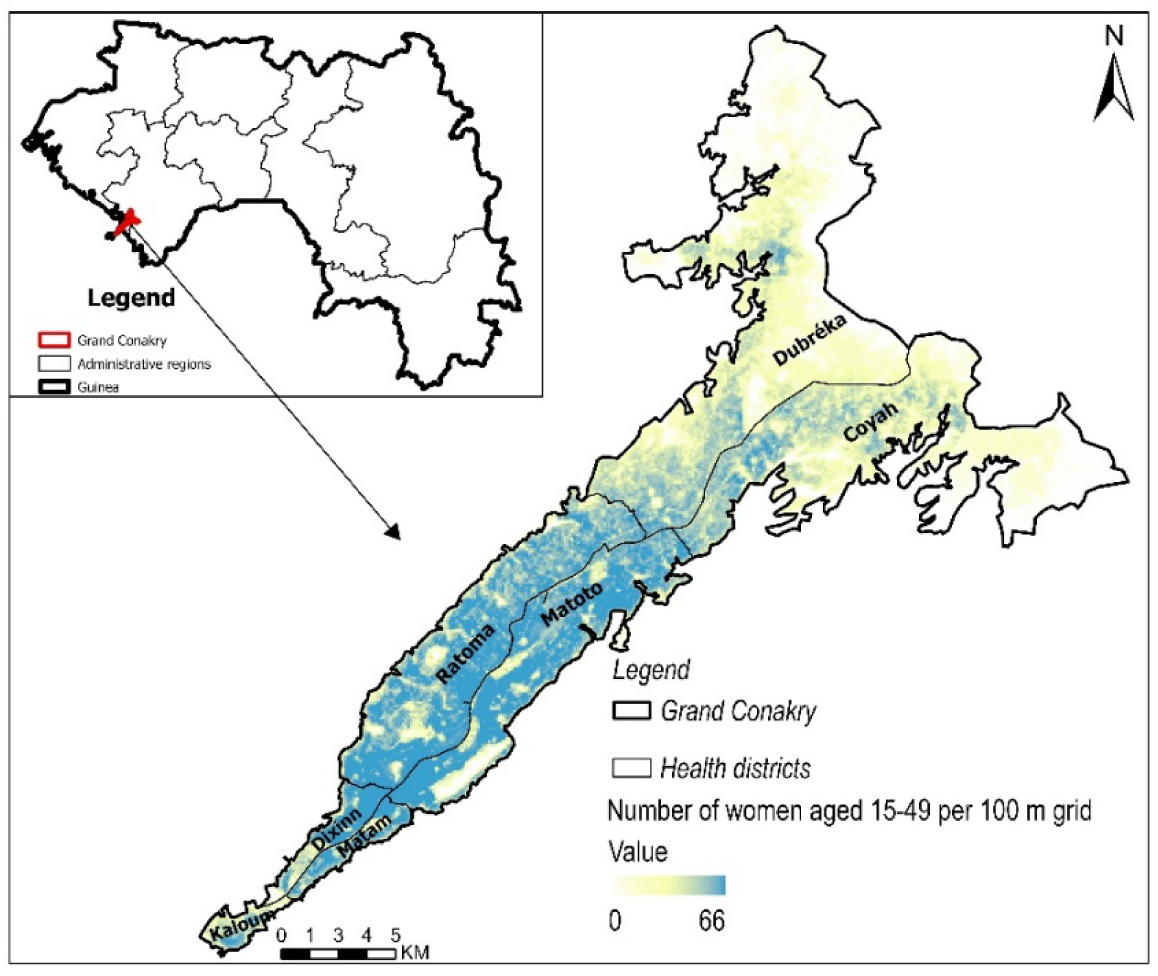
Map showing the population density at the 100 m grid level, ranging from low (0 people per 100 m grid) to high (66 people per 100 m grid) in the urban settings of the seven health districts in Grand Conakry, Guinea. Health district boundaries are delineated in black.^19^

In 2010, Guinea introduced free maternal care in public facilities, including for vaginal and caesarean births. Maternal healthcare service coverage is high in Conakry, with around 90% of births taking place in health facilities, 25% of which were in public hospitals.^24,25^ Maternal and newborn services are provided by public and private sectors. Antenatal care consultations and basic emergency obstetric care (BEmOC) are mainly provided in public facilities (health centres), supported by private not-for-profit facilities. Comprehensive emergency obstetric care (CEmOC) is limited to public hospitals (district, regional and national), and a few private hospitals.^26^

Several initiatives aimed to document and map health facilities and the services they provide in Grand Conakry. These include administrative listings held by the Ministry of Health and its decentralised units (municipal and prefectural health directions (DCS and DPS)), and databases developed as part of vertical programmes such as immunisation programmes and community health activities. According to a study published in 2025 based on a list from the Ministry of Health and Public Hygiene, there were 155 health facilities in Grand Conakry: 58 public and 84 private, including 13 private not-for-profit.^19,27^

### Study design

This was a cross-sectional observational quantitative study aiming to compile a comprehensive geotagged list of public, private-for-profit (PFP) and not-for-profit health facilities operating in Grand Conakry at the time of data collection, with a focus on availability of maternal and newborn health services. The protocol of this study is published elsewhere.^28^

### Sampling and recruitment

We aimed to identify all health facilities in Grand Conakry. We cross-referenced lists of public and private health facilities in Grand Conakry from various departments of the Ministry of Health: The Office of Strategy and Development (BSD/SNISS), the Community Health Department, and the National Directorate of Public and Private Institutions (DNEPP). We compared these lists with those obtained from the seven health districts; resulting in a list of 459 health facilities, which served as the foundation for the census. During fieldwork, we adopted a snowball sampling technique to identify new or missed facilities, by asking staff of visited facilities to indicate at least three functional facilities in their vicinity. If the mentioned facilities were not on the original list, they were added and visited by the research team.

### Data collection

Data were collected in July 2025 using a questionnaire on KoboCollect. Data collection was led by a team of 16 data collectors and supervisors (graduate medical doctors and research assistants) and supported by community health workers. The tool was pre-tested in a health district not included in the study, and the questions were subsequently refined. The questionnaire was structured in two sections: general questions and questions relating to maternal and newborn health services. The first section documented general facility characteristics (*e.g., GPS coordinates, district, name of the facility, sector, type and level of care*, opening times and integration into the district health information system 2 (DHIS2)).These questions were administered to managers, heads of departments or another managerial staff member (their deputy or executive assistant). The second section collected information on the availability of maternal and newborn health services, including opening hours, availability of obstetric and newborn signal functions, out-of-pocket fees charged for care, intra-facility referral linkages. These questions were generally administered either to health facility managers or to senior midwives. When the respondent differed from the one answering section 1, verbal consent was requested from the new respondent. The full questionnaire was published elsewhere.^28^

### Variable Definition and data analysis

Table 1 includes a list of analysed variables and their definitions and categories. Facilities that were closed, non-operational, or refused to participate were excluded from the analysis. The analysis combined quantitative and geospatial descriptive approaches, following the two sections of the questionnaire. The first section focused on sector, type and level of care, and integration into district health information system 2 (DHIS2), stratified by health district. Next, we mapped the coordinates of included facilities, using supplementary geographical data to precisely delineate and adjust the boundaries of health areas (aires de santé) within each district. Last, we described the availability of maternal and newborn healthcare services, particularly BEmONC and CEmONC, receiving and sending obstetric referrals, and fees charged for care, stratifying by sector. The results are presented in as frequencies and percentages, medians and ranges, in tables graphs and maps. Analyses were conducted using Stata version 16 and ArcGIS Pro version 3.7.

**Table 1:** Definitions of the study variables.

| Variable and definition | Categories |
| --- | --- |
| **Health district** | Kaloum, Matam, Dixinn, Ratoma, Matoto, Dubréka, Coyah |
| **Sector** - ownership and management of health facilities | **Public** sector: include all health facilities (from health posts/centres to hospitals), managed and funded by the government under the authority of the Ministry of Health.<br>**Private** sector: comprises all non-governmental, privately funded health facilities, regardless of their legal status, and is divided into:<br>- *For-profit (FP)*: entities providing care to generate a profit, such as clinics and medical practices, often concentrated in urban areas.<br>- *Not-for-profit (NFP)*: organisations, NGOs or faith-based institutions providing care on a non-profit basis, reinvesting their resources in health services and often complementing the public sector. |
| **Level** - health service categorisation in the Guinean context, the national health system is structured into three levels for the public sector. | Within the public sector, 3 levels:<br>• **Primary**: delivered at community health posts/centres, including basic consultations, maternal and child health, vaccinations, and simple treatments. It is also the first contact between local community and the healthcare system.<br>• **Secondary**: provided at district or regional hospitals (HP/HR), including hospitalisation, surgeries, and management of more complex conditions. They represented the first level of referral.<br>• **Tertiary**: offered at national or specialized hospitals, including advanced surgery, intensive care, and management of complex or rare diseases.<br>Within the private sector, three levels:<br>• Private dispensary and health centre (only basic care)<br>• Private cabinet & medical centre (some specialised care services (obstetrics, gynaecology, paediatrics), in addition to primary care)<br>• Private clinics & polyclinics (with a surgical unit)<br>Military health services – includes military and paramilitary separately because although they are public institutions, they do not fall directly under the Ministry of Health. |
| **DHIS2 integration** | Facility reports to the District Health Information System 2 (DHIS2):<br>1) Yes, directly, 2) Yes, via another facility, 3) No, the facility does not report |
| **Services provided in the last 12 months (yes, no)** | *Family planning services (FP)*, Antenatal care (ANC), Childbirth care, Postnatal care (PNC), Care to premature newborns, Vaccination (newborns/babies), *caesarean section*, *blood transfusion* |
| **BEmONC (yes, no)** | Seven functions: Parenteral administration of antibiotics, Parenteral administration of oxytocin, Parenteral administration of magnesium sulphate, Assisted vaginal delivery, Manual removal of placenta, Removal of retained products, Basic newborn resuscitation.<br>Asked whether providers working in the facility had provided/performed the function in the past 12 months. |
| **CEmONC (yes, no)** | BEmONC functions plus: caesarean section, blood transfusion<br>Asked whether providers working in the facility had provided/performed the function in the past 12 months. |
| **Obstetric referrals** | Whether the facility reported receiving (yes, no) or sending (yes, no) referrals of women who are pregnant, in labour, or postpartum. |
| **Fees for vaginal delivery** | 1) Free with free kit, 2) Free, kit not free, 3) Do not know/ no answer (whether the kit of free or not), 4) Not free – fees reported in Guinean francs (GNF) |
| **Fees for delivery by caesarean section** | 1) Free with free kit, 2) Free, kit not free, 3) No answer/Do not know (whether the kit of free or not), 4) Not free – fees in Guinean francs (GNF) |

## RESULTS

### Number of facilities providing healthcare in Grand Conakry

From the consolidated list of health facilities (n=459) obtained from health authorities, we identified, located and visited 231 (50.4%); 228 could not be located, were closed, or not functioning (due to a change of location or name). With the snowballing technique we identified 564 facilities that were not on the health authorities’ list. In total we visited 795 facilities. Of these**, 691** agreed to participate in the study, and 104 were closed (n=63), not operational (n=12), or refused to participate (n=29). Among the facilities that participated in the study**, 331** reported offering childbirth care services and agreed to complete the second section of the questionnaire (Figure 2).

**Figure 2.**
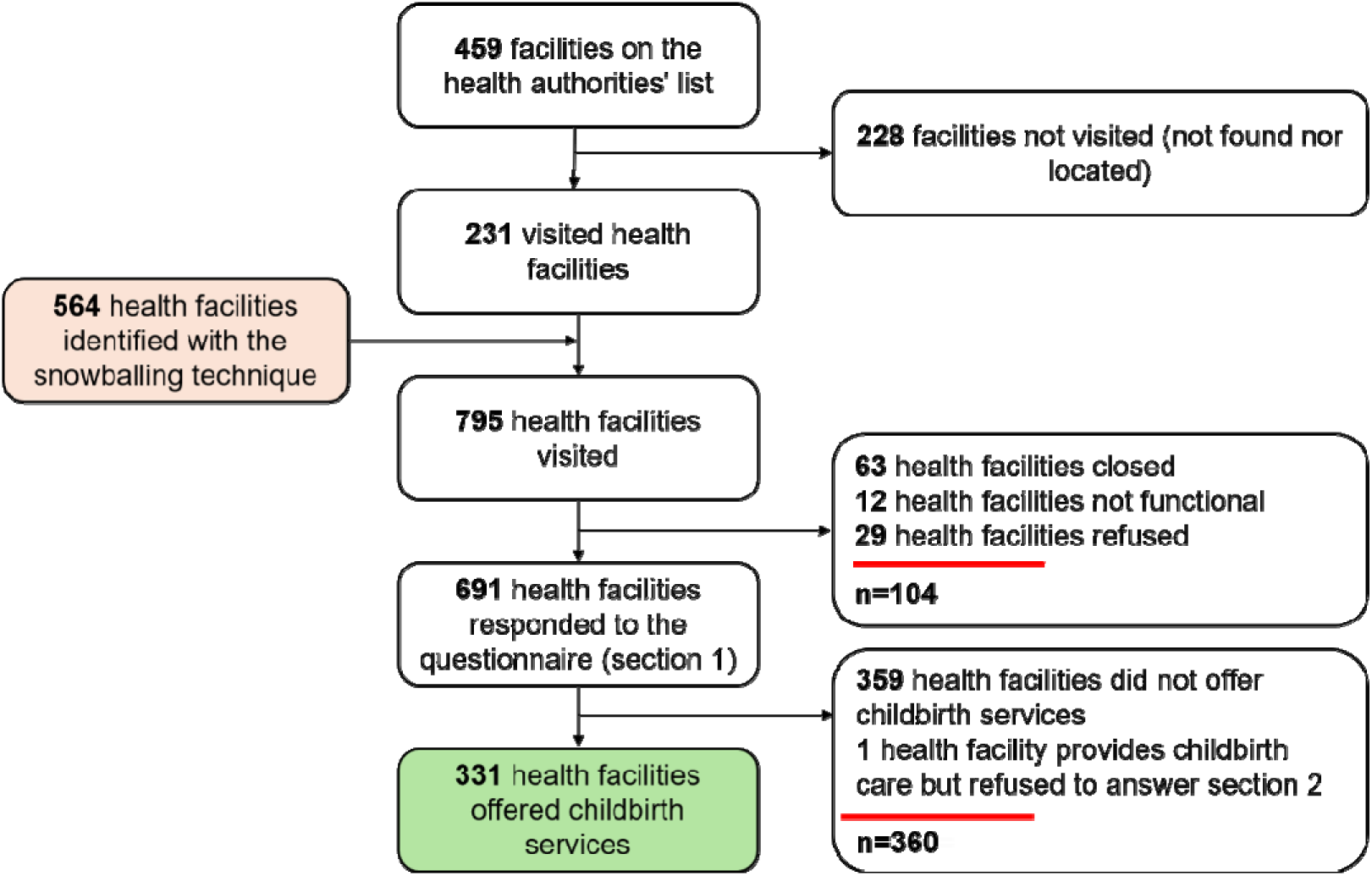
Flowchart of inclusion of health facilities in the census in Grand Conakry, July 2025. **Note:** in this study, a closed facility means that the door was padlocked when team members visited. Most of these were closed following an order of the health authorities due to non-compliance with the required standards.

The map in *Figure 3* shows the geographic distribution of the 691 health facilities in Grand Conakry, spread across 39 health areas covering the seven health districts. Primary facilities constitute most of the sample and are evenly distributed geographically, while most tertiary facilities are clustered in the city centre.

**Figure 3.**
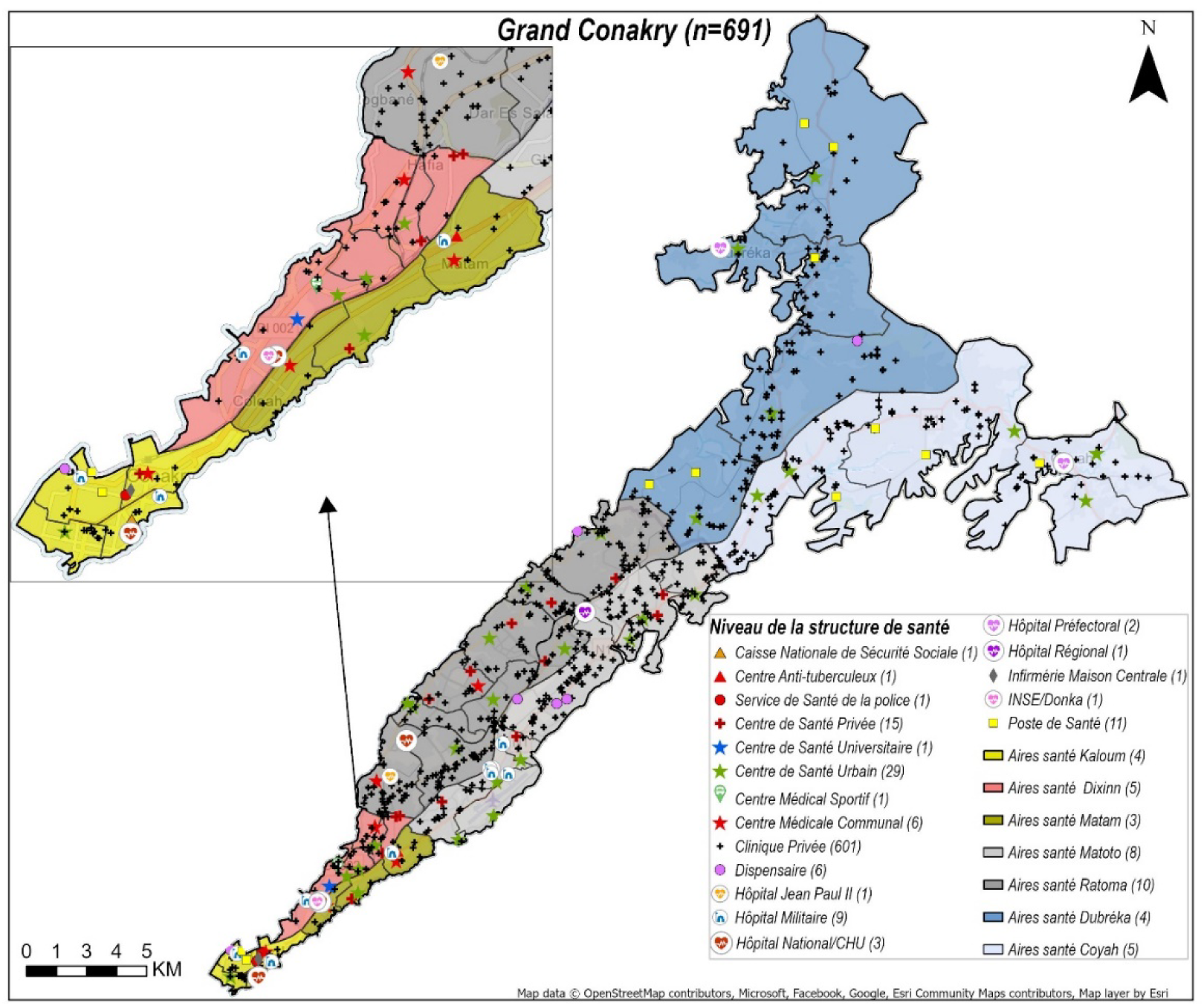
Spatial distribution of 691 health facilities in Grand Conakry

### General characteristics of health facilities

Table 2 shows the characteristics of the 691 facilities that answered the first section of questionnaire. The public sector accounted for 11.1% of facilities (n=77) compared to 88.9% in the private sector (n=614); of which private-for-profit were 74.2% (n=513) and 14.7% (n=101) were private not-for-profit. Ratoma had the highest number of facilities (220 facilities (32%)), of which 94.1% were private. Matam had the lowest number of facilities (n=17, 2.5%), and the highest percentage of public facilities (35.3%). Private clinics and polyclinics were the most common level of facilities, accounting for 55.7% (n=385), followed by private cabinet & medical center, which accounted for 30.4% (n=210). These private health facilities were mainly concentrated in Ratoma, Dubréka Matoto. Public secondary care-level facilities were evenly distributed across the seven health districts.

**Table 2.** General characteristics of health facilities in the Grand Conakry, by district (n=691)

|  | Kaloum<br>n (%) | Matam<br>n (%) | Dixinn<br>n (%) | Ratoma<br>n (%) | Matoto<br>n (%) | Dubréka<br>n (%) | Coyah<br>n (%) | Total<br>n (%) |
| --- | --- | --- | --- | --- | --- | --- | --- | --- |
| **Total (facilities) - row %** | **32 (4.6)** | **17 (2.5)** | **40 (5.8)** | **220 (31.8)** | **158 (22.9)** | **129 (18.7)** | **95 (13.7)** | **691 (100)** |
| **Sector (column %)** |  |  |  |  |  |  |  |  |
| *Public* | 11 (34.4) | 6 (35.3) | 9 (22.5) | 13 (5.9) | 16 (10.1) | 11 (8.5) | 11 (8.6) | 77 (11.1) |
| *Private for-profit* | 20 (62.5) | 10 (58.8) | 26 (65.0) | 184 (83.6) | 128 (81.0) | 82 (63.6) | 63 (66.3) | 513 (74.2) |
| *Private not-for-profit* | 1 (3.1) | 1 (5.9) | 5 (12.5) | 23 (10.5) | 14 (8.9) | 36 (27.9) | 21 (22.1) | 101 (14.7) |
| **Type of health facility (column %)** |  |  |  |  |  |  |  |  |
| *Public primary level* | 6 (18.7) | 2 (11.8) | 5 (12.5) | 9 (4.1) | 10 (6.3) | 10 (7.7) | 10 (10.5) | 52 (7.5) |
| *Public secondary level* | 2 (6.2) | 2 (11.8) | 1(2.5) | 3 (1.4) | 1 (0.6) | 1 (0.8) | 1 (1.0) | 11 (1.6) |
| *Public tertiary level* | 1 (3.1) | 1 (5.9) | 2 (5.0) | 1 (0.4) | 0 (0.0) | 0 (0.0) | 0 (0.0) | 5 (0.7) |
| *Private dispensary and health centre* | 1 (6.2) | 1 (5.9) | 2 (5.0) | 7 (3.2) | 6 (3.8) | 1 (0.8) | 0 (0.0) | 19 (2.8) |
| *Private cabinet & medical centre* | 11 (34.4) | 0 (0.0) | 17 (42.5) | 63 (28.6) | 48 (30.4) | 26 (20.1) | 45 (47.4) | 210 (30.4) |
| *Private clinics & polyclinics* | 8 (25.0) | 10 (58.8) | 12 (30.0) | 137 (62.3) | 88 (55.7) | 91 (70.6) | 39 (41.0) | 385 (55.7) |
| *Military health services* | 2 (3.2) | 1 (5.9) | 1 (2.5) | 0 (0.0) | 5 (3.2) | 0 (0.0) | 0 (0.0) | 9 (1.3) |

Table 3 shows the integration of health facilities into DHIS2, by sector. More than half of health facilities (n=433; 62.7%) reported that they did not transmit routine data to the DHIS2. Most public sector facilities (n=66/77, 85.7%) reported their data to the DHIS2, either directly (n=54, 70.1%) or via another facility (n=12, 15.6%). Data transmission into DHIS2 was limited in the private sector, both among for-profit (n=159/513, 31%) and not-for-profit facilities (n=33/101, 32.7%).

**Table 3.** Health facilities integration in the DHIS2, by sector (n=691)

|  | Public sector<br>n (%) | Private sector |  | Total<br>n (%) |
| --- | --- | --- | --- | --- |
| **DHIS2 integration (column %)** |  | For-profit n (%) | Not-for-profit n (%) |  |
| *Yes, directly* | 54 (70.1) | 116 (22.6) | 19 (18.8) | 189 (27.3) |
| *Yes, through another facility* | 12 (15.6) | 43 (8.4) | 14 (13.9) | 69 (10.0) |
| *No* | 11 (14.3) | 354 (69.0) | 68 (67.3) | 433 (62.7) |

### Services provided by health facilities

Table 4 shows the distribution of the healthcare services provided by health facilities in Grand Conakry. Approximately half offered family planning services, antenatal care and postnatal care services. Childbirth care services were reported being offered by 332 facilities (48.7%). In contrast, care for premature newborns was offered by 40 facilities (5.8%), approximately evenly distributed by district. Child vaccination services were offered by 115 of 691 the facilities (17%).

**Table 4.** Description of services provided in the last 12 months by health facilities in Grand Conakry, by health district (n = 691)

|  | Kaloum<br>n (%) | Matam<br>n (%) | Dixinn<br>n (%) | Ratoma<br>n (%) | Matoto<br>n (%) | Dubréka<br>n (%) | Coyah<br>n (%) | Total<br>n (%) |
| --- | --- | --- | --- | --- | --- | --- | --- | --- |
| **Total (facilities)** | **32** | **17** | **40** | **220** | **158** | **129** | **95** | **691** |
| *Family planning services* | 7 (21.9) | 7 (41.2) | 23 (57.5) | 117 (53.2) | 89 (56.4) | 80 (62.0) | 57 (60.0) | 380 (55.0) |
| *Antenatal care* | 9 (28.1) | 7 (41.2) | 12 (30.0) | 117 (53.2) | 87 (55.1) | 78 (60.5) | 41 (43.2) | 351 (50.8) |
| *Childbirth care* | 8 (25.0) | 7 (41.2) | 11 (27.5) | 105 (47.7) | 78 (49.4) | 76 (58.9) | 47 (49.5) | 332 (48.0) |
| *Postnatal care* | 7 (21.9) | 7 (41.2) | 15 (37.5) | 105 (47.7) | 76 (48.1) | 84 (65.1) | 56 (58.9) | 350 (50.7) |
| *Care for premature newborns* | 2 (6.3) | 2 (11.8) | 5 (12.5) | 16 (7.23) | 7 (4.4) | 4 (3.1) | 4 (4.2) | 40 (5.8) |
| *Childhood vaccinations* | 6 (18.8) | 5 (29.4) | 10 (25.0) | 40 (18.2) | 28 (17.7) | 16 (12.4) | 10 (10.5) | 115 (16.6) |

### Facilities offering childbirth care services

Table 5 shows the availability and distribution of emergency obstetric signal functions. This section of the questionnaire was asked only to facilities that declared offering childbirth care and agreed to continue to answer the second section of the questionnaire (n=331). Most of the 331 facilities (n=294, 88.8%,) provided continuous service 24 hours a day, 7 days a week, including all (n=53) public health facilities providing childbirth services.

**Table 5.** Description of signal functions provided in the 12 months preceding the census by health facilities offering childbirth care in the Grand Conakry, by sector (n=331)

|  | Public<br>n (%) | Private for-profit<br>n (%) | Private not-for-profit<br>n (%) | Total<br>n (%) |
| --- | --- | --- | --- | --- |
| **TOTAL** | **53** | **224** | **54** | **331** |
| **BEmONC signal functions** |  |  |  |  |
| *Parenteral administration of antibiotics* | 50 (94.3) | 213 (95.1) | 50 (92.6) | 313 (94.6) |
| *Parenteral administration of oxytocin* | 50 (94.3) | 212 (94.6) | 49 (90.7) | 311 (94.0) |
| *Parenteral administration of magnesium sulphate* | 28 (52.8) | 91 (40.6) | 15 (27.8) | 134 (40.5) |
| *Assisted vaginal delivery* | 30 (56.6) | 60 (26.8) | 4 (7.4) | 94 (28.4) |
| *Manual removal of placenta* | 47 (88.7) | 161 (71.8) | 39 (72.2) | 249 (75.2) |
| *Removal of retained products* | 47 (88.7) | 179 (82.1) | 47 (87.0) | 275 (83.1) |
| *Basic newborn resuscitation* | 49 (92.5) | 165 (73.6) | 35 (64.8) | 249 (75.2) |
| **All 7 BEmONC signal functions** | 23 (43.4) | 71 (32.7) | 12 (22.2) | 106 (32.0) |
| **CEmONC signal functions** |  |  |  |  |
| *Caesarean section* | 11 (20.8) | 103 (46.0) | 18 (33.3) | 132 (39.9) |
| *Blood transfusion* | 11 (20.8) | 63 (28.1) | 13 (24.1) | 87 (26.3) |
| **All 9 CEmONC signal functions** | 6 (11.3) | 9 (4.0) | 1 (1.8) | 16 (4.8) |
| **Referrals during pregnancy, labour, or postpartum** |  |  |  |  |
| *Receives referrals* | 26 (49.1) | 91 (40.4) | 20 (37.0) | 137 (41.3) |
| *Sends referrals* | 50 (94.3) | 200 (88.9) | 52 (96.3) | 302 (91.0) |
| **Open 24-hours, 7-days a week** |  |  |  |  |
| **Yes** | 53 (100.0) | 195 (87.1) | 46 (86.2) | 294 (88.8) |

Individual BEmONC functions, such as parenteral administration of antibiotics and oxytocin, were available in more than 90% of facilities in both sectors (public and private) followed by removal of retained products and manual removal of the placenta. However, magnesium sulphate administration and assisted vaginal deliveries were provided by less than half of the facilities, and more frequently available in the public sector compared to the private sector. A total of 132 facilities (40%) declared performing caesarean sections, 121 private and 11 public. In addition, 87 (26%) facilities reported providing blood transfusion, most of them were private-for-profit. 106 of the 331 health facilities providing childbirth care (32%) reported providing all seven BEmONC functions, 23 in the public sector. 16 health facilities (5%) provided all nine CEmONC functions; 6 were public. Regarding obstetric referrals, 302 (91%) facilities offering childbirth care reported that they referred women during pregnancy, labour or the postpartum period to other facilities, and 137 (41%) stated that they receive referred obstetric patients.

### Fees for childbirth care services

Table 6 shows the fees facilities reported charging for vaginal and caesarean births. Among facilities offering vaginal births (n=331), 13 facilities (3.9%) reported providing free deliveries with a free kit; all public. About 60% of public facilities responded “no-answer/don’t know” about whether they charge fees for vaginal births. Among facilities which reported the amount they charge for vaginal births, the median ranged from 50,000 GNF (6 USD) in the public sector to 250,000 (29 USD) in the private for-profit sector.

**Table 6.**
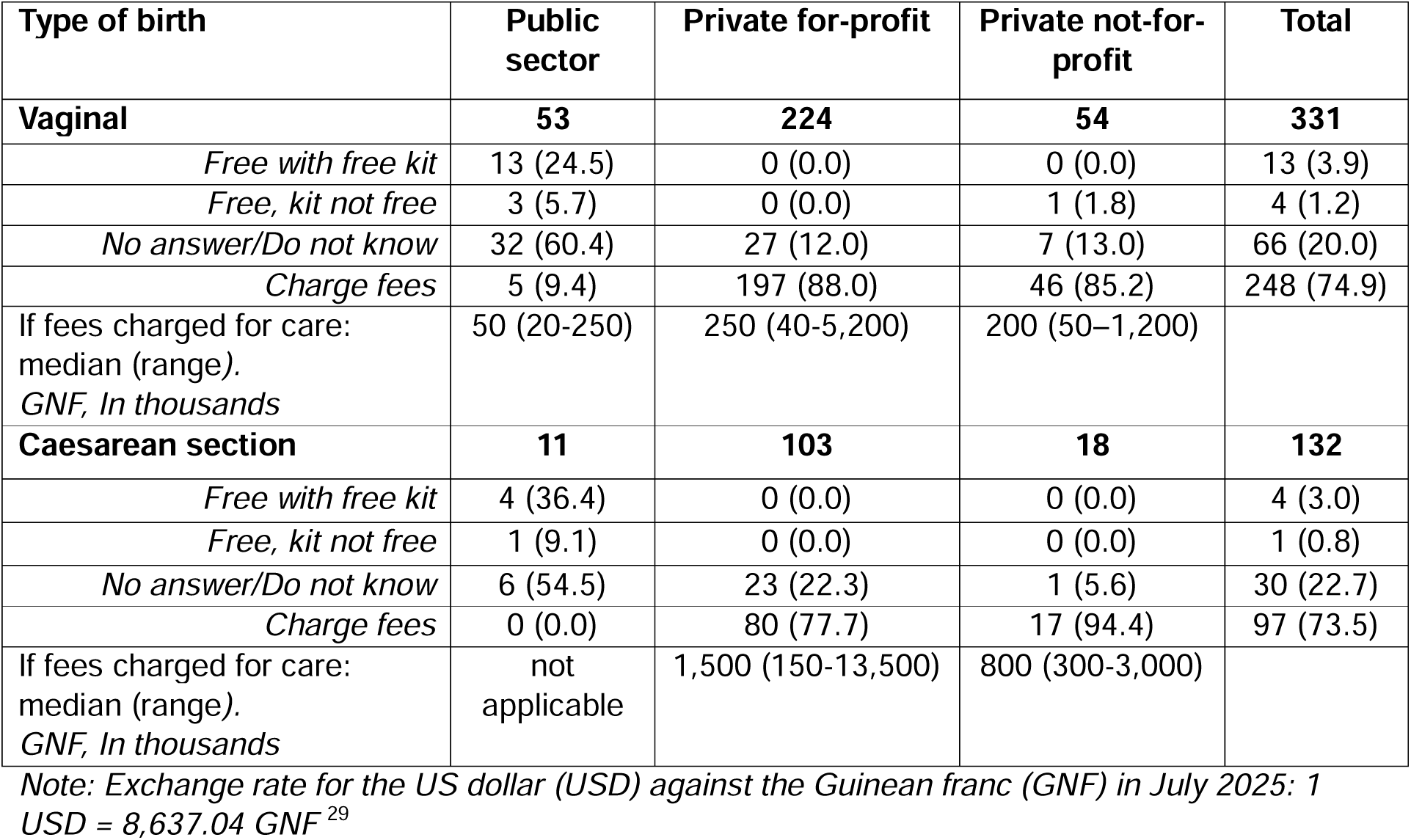
Out-of-pocket fees for of childbirth care by mode of birth of access to childbirth kits in health facilities offering childbirth care in Grand Conakry, by sector.

| Type of birth | Public sector | Private for-profit | Private not-for-profit | Total |
| --- | --- | --- | --- | --- |
| **Vaginal** | **53** | **224** | **54** | **331** |
| *Free with free kit* | 13 (24.5) | 0 (0.0) | 0 (0.0) | 13 (3.9) |
| *Free, kit not free* | 3 (5.7) | 0 (0.0) | 1 (1.8) | 4 (1.2) |
| *No answer/Do not know* | 32 (60.4) | 27 (12.0) | 7 (13.0) | 66 (20.0) |
| *Charge fees* | 5 (9.4) | 197 (88.0) | 46 (85.2) | 248 (74.9) |
| If fees charged for care:<br>median (range).<br>*GNF, In thousands* | 50 (20-250) | 250 (40-5,200) | 200 (50–1,200) |  |
| **Caesarean section** | **11** | **103** | **18** | **132** |
| *Free with free kit* | 4 (36.4) | 0 (0.0) | 0 (0.0) | 4 (3.0) |
| *Free, kit not free* | 1 (9.1) | 0 (0.0) | 0 (0.0) | 1 (0.8) |
| *No answer/Do not know* | 6 (54.5) | 23 (22.3) | 1 (5.6) | 30 (22.7) |
| *Charge fees* | 0 (0.0) | 80 (77.7) | 17 (94.4) | 97 (73.5) |
| If fees charged for care:<br>median (range).<br>*GNF, In thousands* | not applicable | 1,500 (150-13,500) | 800 (300-3,000) |  |

*Note: Exchange rate for the US dollar (USD) against the Guinean franc (GNF) in July 2025: 1 USD = 8,637.04 GNF*<sup>29</sup>

Among the 132 facilities providing caesarean section, four (3.0%) reported offering free caesarean sections with a free kit, all public. No public facilities reported charging fees for caesarean sections, although 6 of the 11 did not provide an answer. Among the facilities which reported the amount they charged for caesarean sections, the median ranged from 800,000 GNF (93 USD) in the private not-for-profit sector to 1,500,000 (174 USD) in the for-profit sector.

## DISCUSSION

This study used data from a comprehensive census to describe the characteristics of health facilities in Grand Conakry in 2025, with a particular focus on the availability of maternal and newborn healthcare services. We focus this discussion on three key points. First, the discrepancy between the number and type of facilities between authorities’ and our lists, a result of weak administrative mechanisms for updating health facility lists and resulting challenges for regulating public and private healthcare provision. Second, we discuss the gaps in the availability of emergency obstetric and newborn services. Third, we reflect on the findings of out-of-pocket cost of childbirth care.

First, Grand Conakry had 691 operational health facilities in July 2025; 564 more than on the health authorities’ lists. In addition, of the 459 facilities on the official list, half were not found during fieldwork, either because they were closed or because they could not be located. The majority of the facilities which were successfully identified in this census fieldwork were in the private sector.

Data quality in the DHIS2 relies on an up-to-date and comprehensive list of health facilities, in order to serve as the reference framework for the health system.^22,30,31^ Our findings suggest that administrative processes of updating health facility lists struggle to keep pace with the urban expansion of Grand Conakry over time. In fact, the opening, reopening, closure and changes in status or location of facilities are not systematically reported, resulting in incomplete or outdated lists. This situation additionally reflects a governance issue. According to the World Health Organization, a national register of health facilities (Master Facility List) must be comprehensive, regularly updated and governed by robust institutional mechanisms to support the planning, coordination and management of healthcare services.^32^ It should be emphasised that this shortcoming particularly affects the private sector, whose growth often outpaces administrative regulatory capacities, especially in major cities of sub-Saharan Africa,^19,22^ leading to the under-representation of private healthcare provision in national databases.

Our findings also show that more than three-quarters of private health facilities (both for-profit and not-for-profit) do not submit their data to DHIS2, far below the proportion of public sector facilities doing so. This under-representation of the private sector in national health information systems, is well documented in sub-Saharan Africa.^19^ In Grand Conakry, this situation could be explained by a collaboration that has historically been centred on the public sector, with systems for supervision, regulation and performance monitoring having been developed primarily for public facilities. This discrepancy between administrative data and the ground truth reduces health authorities’ capacity to gain a comprehensive overview of healthcare provision. Such an overview is necessary to guide planning, resource allocation and decision-making, to and enables the health system to respond effectively to the population’s needs, particularly during health emergencies, to which cities are particularly susceptible.

We also found a marked concentration of facilities in the districts of Ratoma and Matoto, the most densely populated areas in Grand Conakry. More than 90% of the facilities identified were private, particularly in Ratoma and Matoto. This pattern could be explained by rapid urbanisation linked to population growth in these districts, which has fostered the expansion of the private health sector. Facilities in the public sector are clearly not meeting the growing demand for healthcare services, and this could be related to both the volume of the demand, and other attributes (accessibility, acceptability, quality and affordability). ^19,22^ This situation is comparable to that observed in other sub-Saharan African cities, such as Lubumbashi and Douala, where private health facilities constitute most facilities.^33,34^ However, this does not necessarily mean that they provide the majority of services, as many of these facilities operate on a relatively small scale and do not provide advanced care.^33^

Second, regarding the provision of maternal healthcare, our findings show a troubling paradox within health facilities. Among the 331 facilities providing childbirth care, only one third offer complete BEmONC functionality. This means that most facilities are technical incapable of providing comprehensive care for obstetric and newborn complications, which reduces the chances of positive outcomes.^6,35^ For example, assisted vaginal deliveries(basic and routine BEmONC procedures), are available about a quarter of facilities. This calls into question the quality of care and responsiveness of teams assisting childbirth care. Main barriers to this functionality are shortages in qualified healthcare personnel with training, shortage of functional equipment (e.g. vacuum extractors and/or forceps) and medico-legal implications (e.g., liability in case of complications, fear of legal action and sanctions, or lack of guidelines governing use). This underscores the shortage ineffectiveness of current mechanisms for regulating and supervising childbirth care provision, particularly in highly privatised context.^36,37^ In Grand Conakry, only 16 out of the 331 facilities caring for women during childbirth offered all nine CEmONC functions, of which six were in the public sector. This concentration of care for complications could lead to overcrowded CEmONC facilities, prolong the time taken to provide care and, consequently increasing the risk of adverse outcomes for women and newborns, particularly when rapid intervention is critical.

Third, while the policy clearly states that childbirth care and delivery kits are to be provided free-of-charge in public facilities, this was only reported by one-quarter of them. More than half of public facilities providing childbirth care avoided responding to this question; seemingly because they are aware that care is to be provided for free but effectively, is not. In our census, only 13 facilities in Grand Conakry reported providing free vaginal births and four free caesarean section care, including the delivery kit. Yet, this number is likely overestimating the true availability of completely free care which is (no cash or gifts, whether formal or informal, requested by health facilities or individual health workers). Thus, the user-fee removal policy, which was intended to improve equity by enhancing financial access to care for all, remains largely theoretical in this context.^38^ The policy faces challenges ineffective implementation and the sustainability of the financial protection it is supposed to offer, especially for low-income households, which continue to face indirect costs such as transportation, unsubsidised medication, additional tests and informal fees.^39–41^ This issue extends to other services, as many adolescents and young people remain unable to access contraceptive services due to their financial dependence on adults. As long as free care is not available and sustainable, the most vulnerable groups will continue to forgo healthcare services, delay seeking care, or incur catastrophic health expenditures.^42^

This finding is more worrying given that the healthcare landscape in Grand Conakry is numerically dominated by private-for-profit facilities, who rely on out-of-pocket payments. When the few public facilities offering emergency obstetric and newborn care do not offer free care, socio-economically disadvantaged women have few alternatives. This situation reduces the redistributive effect of policies guaranteeing free care and risks exacerbating inequalities in access to maternal and newborn care, particularly in an urban context. It also highlights the limitations of collaboration between the public and private sectors, where a lack of coordination undermines the objective of equitable access to care.^21,43^ The rapidly growing landscape of care provision does not, on its own, guarantee equitable access to maternal and newborn care. In the absence of effective financial protection, regulation of care quality, and better integration of the private sector into public policies, economic barriers will continue to limit access to care for the disadvantaged populations in Grand Conakry.

## STRENGTHS AND LIMITATIONS OF THE STUDY

A key strength of this study is its census-based approach, well-suited to providing a comprehensive overview of healthcare availability. Where existing data was fragmented and outdated, this method provided a more complete picture of the urban healthcare landscape. Furthermore, the study combines descriptive and geospatial analyses to examine facilities’ geographical distribution and characteristics. Including the public and private sectors is another major strength. Finally, analysing BEmONC and CEmONC functions’ availability enables us to examine facilities’ operational capacity.

This study has certain limitations. While this was a census, there is a risk of selection bias because of refusal to participate. It is possible that facilities which refused are likely to suffer from issues such as unavailability of key service packages and violation of the user fee removal policy. Additionally, some of the information is based on statements from facility managers, which carries information bias.

## CONCLUSION

00000Despite the high number of health facilities in Grand Conakry, this study highlights important challenges limiting access to quality maternal and newborn care. The lack of up-to-date inventory of health facilities undermine effective health system governance, constrain evidence-based planning, and limit the integration of the predominant private sector into health system management. At the same time, important gaps remain in the availability and quality of emergency obstetric and newborn care. Persistent out-of-pocket payments for childbirth services continue to create financial barriers despite national user-fee-removal policies. This study demonstrates that improving maternal and newborn health in rapidly urbanizing Grand Conakry requires expanding service availability, strengthening health system governance, maintaining accurate and up-to-date information on health facilities, ensuring effective oversight of public and private sectors, improving the quality of emergency obstetric and newborn care, and reducing the financial burden on women seeking childbirth care.

## Supporting information

STROBE-checklist-v4-cross-sectional 1

## Acknowledgements

The authors would like to express their sincere thanks to all the institutions that supported this research and to the study participants for the valuable time they devoted to answer the questionnaire and their willingness to take part. We are equally grateful to the data collectors for their dedication and commitment throughout the data collection process for this study. he authors also wish to express their sincere appreciation to Ms. Angèle Soua KOLIE for her constant administrative and financial support throughout this study.

## Ethics

the study was reviewed and approved by the Institute of Tropical Medicine (ITM) in Antwerp Ethics Committee and the Health Research Ethics Committee (CNERS) of Guinea, (references: IRB/1877/25 and 098/CNERS/25). Administrative authorisations were obtained from health authorities and health facility managers, and verbal consent was obtained from respondents. The project was supported by the Ministry of Health (through its departments) and the national office of Private, confessional and associative health facilities in Guinea (Réseau National des Organisations Sanitaires Privées, Associatives et Confessionnel de Guinée: RENOSPAC).

## Data Availability

the dataset will be published later in open access, following final approval from the relevant health authorities, in accordance with the applicable regulatory requirements. Data will be made available upon reasonable request.

## Authors’ contributions

ND, AS, RO, MG, PM, AD, and LB conceptualized the research idea and contributed to the development of the study design and protocol. ND, AS, RO, and MG supervised/coordinated data collection. JGD, MKK, PK, MS, and HFS conducted, provided supervision and oversight of the data collection process. SD, AKN, TMM, HM, DFK, FC, FBC, JDS, EG, AS, HS, BS, FD, AB, SDS, SC, FS, JPL, MS, ASS, BC, AC, SNC, and MBB contributed to the refinement of the study protocol and the coordination and management of fieldwork activities during data collection. ND and AS performed data cleaning and database management. ND analysed the data and drafted the first version of the manuscript. AS, and LB contributed to the interpretation of the findings. PK developed the study map, under the supervision of MG and PM, with contributions from AS, HS, BS, FD, AB, SDS, SC, FS, JPL, MS, ASS, BC, AC, and SNC for validation purposes. ND, AS, RO, PM, MG, HM, TMM, SS and PK reviewed and edited drafts of the manuscript. AS, AD, and LB critically reviewed the manuscript and provided substantial intellectual input. All authors read and approved the final version of the manuscript and agree to be accountable for all aspects of the work.

## Declaration of generative AI use

the authors declare that they have used a generative artificial intelligence tool to improve the linguistic and editorial quality of the manuscript, particularly for rephrasing, grammatical correction and enhancing the clarity of the text. However, a thorough review and verification of the entire content of the manuscript were carried out prior to journal submission, and the authors assume full responsibility for it.

## Biographical note

this paper was produced within UrbanBirth Collective, a multidisciplinary network bringing together researchers, healthcare professionals, urban planners and decision-makers committed to improving maternal and newborn health in cities across sub-Saharan Africa. The Collective conducts research on urban health systems, care pathways, healthcare provision, and inequities in service access to generate evidence that supports context-specific strategies for improving maternal and newborn health in urban settings. It produces also evidence to inform public policy and practice, with a view to creating healthier, more resilient and more equitable cities.

